# Assessment of Vaccine Managers’ Knowledge and Observance of Vaccination Norms and Standards in an Urban Health District of Yaounde (Cameroon) during the COVID-19 Pandemic

**DOI:** 10.1101/2024.11.07.24316922

**Authors:** Fabrice Zobel Lekeumo Cheuyem, Adidja Amani, Brian Ngongheh Ajong, Raissa Katy Noa Otsali, Ariane Nouko, Edwige Omona Guissana, Christelle Sandrine Ngos, Adama Mohamadou, Guy Stephane Nloga, Michel Franck Edzamba, Esther Andriane Bitye Bi Mvondo, Larissa Linda Eyenga Ntsek, Christian Mouangue, Florence Kissougle Nkongo

## Abstract

**Background:** Routine childhood immunization remains a highly cost-effective public health intervention. Global immunization efforts have increased the coverage of proxy antigens in recent decades. However, annual reports indicated a progressive decline in immunization coverage in Yaounde. This study aimed to identify knowledge gaps and organizational deficiencies in the implementation of routine immunization in health facilities (HFs) in the Efoulan Health District (HD) of Yaounde.

**Methods:** A descriptive cross-sectional study was conducted in Efoulan, an urban HD of Yaounde, the capital of Cameroon, in May 2022. A self-administered questionnaire was used to collect knowledge data from consenting HCWs. An observation grid was used to assess the implementation of some relevant national EPI guidelines and recommendations.

**Results:** A total of 46 HCWs providing vaccinations in the nine health areas of HDs were enrolled in the study. Almost all participants (93.5%) were unable to name all the diseases that the pentavalent vaccine protects against. More than half of the respondents (59%) were aware of the 15-month booster dose of the measles-rubella (MR) vaccine included in the EPI. Half of the respondents were aware of the type of antigen contained in the BCG vaccine (52%). More than two-thirds of the respondents (72%) had not identified the 6-month MR vaccine as recommended without risk for HIV+ children. Most of the respondents were not aware of the management of opened nonlyophilized vials (50%), calculation of the wastage factor (89%) or vaccine requirements (98%). Almost all the respondents (87%) had poor knowledge of vaccination norms and standards. Most of the visited HFs did not follow the monthly EPI monitoring curve (87%), did not store vaccines according to standards (80%), or had an EPI case reporting form (93%). Most of the consultants in visited HFs were not familiar with the case definitions of common vaccine-preventable diseases.

**Conclusions:** The implementation of routine immunization remained poor in most of the HFs in Efoulan HDs because of the lack of knowledge of the EPI norms and standards of the staff. There is a need to strengthen the capacities of HCWs through ongoing training and supervision.

## Background

Routine childhood immunization remains a highly cost-effective public health intervention [1]. Immunization has contributed to significant reductions in morbidity and mortality from vaccine-preventable diseases (VPDs) worldwide [2]. Global immunization efforts have saved approximately 154 million lives over the past 50 years, or approximately 6 lives every minute every year. Most of these lives saved were infants (101 million) [2,3]. Global immunization efforts have increased the coverage of the third dose of the diphtheria‒tetanus‒pertussis (DTP3) vaccine from near zero in 1974 to an impressive 84% today. This progress, which reached a high level of 86% in 2019 before the COVID-19 pandemic, demonstrates the effectiveness of global vaccine programs [4].

Vaccination activities for children aged 0–11 months and pregnant women, led by the Expanded Programme on Immunization (EPI) of the Ministry of Public Health (MINSANTE), are a priority objective in Cameroon’s health strategy [5]. The Efoulan Health District (HD) of Yaounde, with its 54 vaccination centers spread across all health sectors, regularly performs routine and mass vaccination campaigns to meet the targets set by health authorities [6].

However, in recent years, annual reports have indicated a progressive decline in vaccination coverage in Efoulan HD. This was characterized by a decrease in pentavalent vaccine (penta) 3 coverage from 85.8% to 72% between 2020 and 2021 (a decrease of 15.4%) and an increase in penta 1--3 specific drop-out rates from -1.5% to 1.8% [7,8].

In addition, in resource-limited countries such as Cameroon, the emergence of coronavirus disease 2019 (COVID-19) has had a significant effect on the health system and disrupted the implementation of EPI activities by diverting attention and health resources to the COVID-19 response [9,10].

The reports of the coordination meetings with EPI focal points, HD officers and health facility (HF) representatives highlighted several challenges, including COVID-19-related constraints leading to lower uptake of immunization services by parents, failure of technical and organizational components, and quality and management issues related to human resources in health facilities, especially in health facilities identified as having problems in the HD.

In addition, data management issues related to EPI inputs were identified, motivating an operational research study to understand the root causes and promote evidence-based decision-making. This study was designed to identify knowledge gaps among immunization stakeholders and describe the organizational pattern in the implementation of routine vaccination in problematic health facilities in the Efoulan HD of Yaounde.

## Methods

### Study Type

An institution-based cross-sectional study was conducted in the Efoulan HD

### Study period

The study was conducted throughout the month of May 2022.

### Study Site

The Efoulan HD is one of the 32 districts of the Central Region. It comprises nine health areas (eight urban areas and one rural area). The population in 2021 was estimated at 453,047 inhabitants covering an area of over 67 km² (density of 6,761 inhabitants per km²). The HD includes 117 health facilities, including seven public, three denominational and 107 private facilities (Figure 1).

**Fig. 1.**
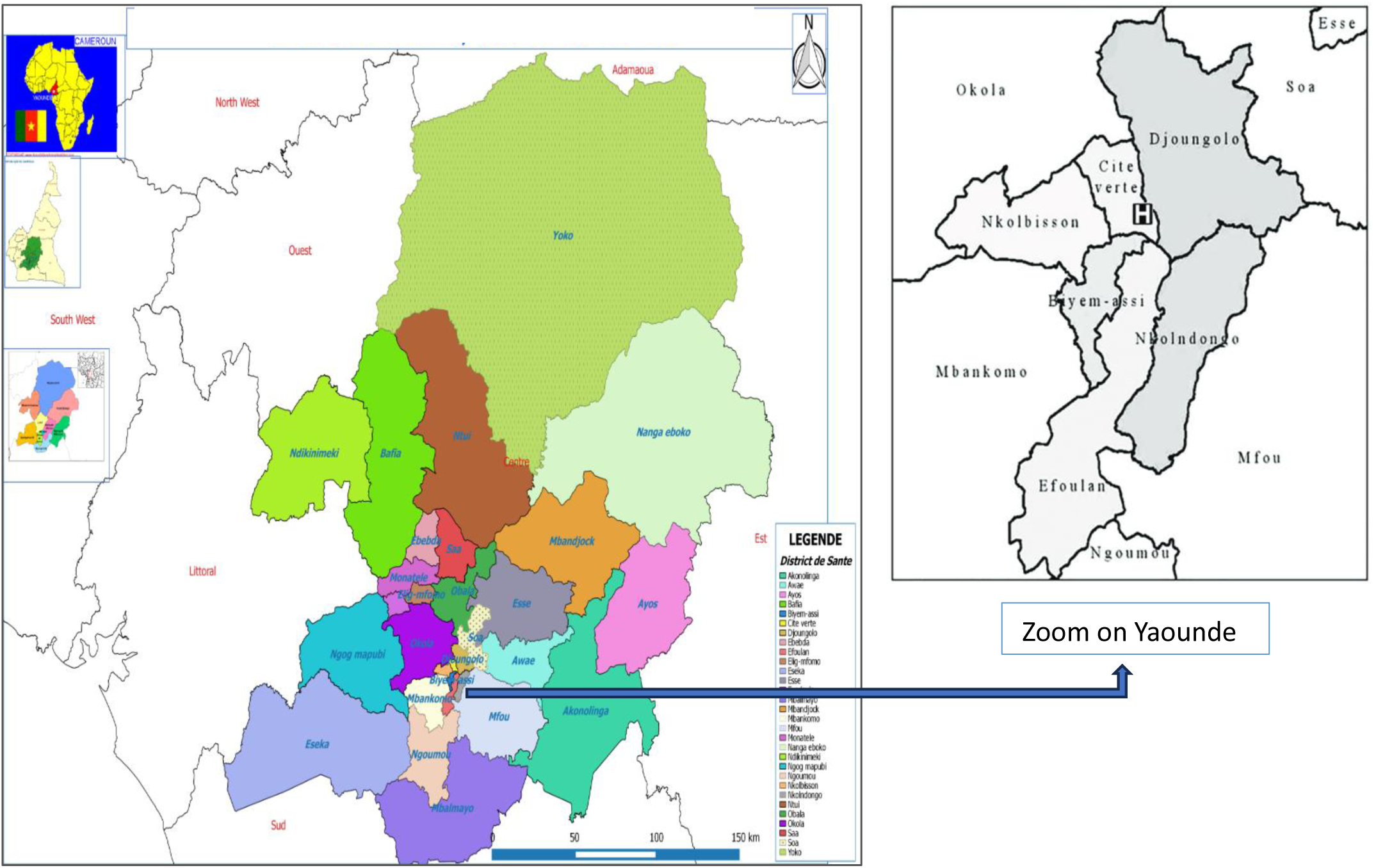
Localization of Efoulan among the Yaounde Health Districts [11].

### Study population

Health workers (HCWs) vaccinating the Efoulan HD and problematic health facilities of the HD.

### Study Participants

HCWs in a HF who were vaccinated with Efoulan HD and who provided informed consent were included.

### Sample size and sampling technique

To ensure the representativeness of all nine health areas, HCWs were enrolled via a purposive sampling technique. For observational analysis, 15 HFs identified as problematic were visited (one public and 14 privates).

### Data collection tool and procedures

Participants were contacted during coordination meetings and field visits. A pretested and self-administered questionnaire of 28 questions was administered, covering sociodemographic information, knowledge of priority targets, vaccination schedules, techniques and input management. An observation grid of 20 preregistered questions on Google Forms was used to collect observational data from 15 sampled health facilities.

### Data processing and analysis

Each participant was assigned a score on the basis of the knowledge assessment (16 questions, 1 point each). The results were then classified into three categories: good (≥75%), average (50–74%), and poor (<50%). [12]. The data were entered into Google Forms, extracted into Microsoft Office Excel 2016 and analyzed via IBM SPSS version 26. Charts were generated via Microsoft Office Excel 2016. For cross-tabulations, proportions were compared via Fisher’s exact tests. A *p* value<0.05 was considered statistically significant.

### Operational Definition

Problematic HFs were described as those with poor performance in implementing immunization programs (low vaccination coverage for penta 3 [<50%] and poor data management in terms of timeliness, completeness and coherence of data recorded in DHIS2) during the year 2021 and who had difficulty mitigating challenges after HD supervision.

## Results

### Study participant profile

A total of 46 HCWs from HFs providing vaccinations in the nine health areas of HDs were enrolled in the study. They were aged 27-53 years, with half of the participants aged between 27 and 36 years. Most of the participants were female (84.8%) and had higher educational levels (63%) (Table 1).

**Table 1.** Sociodemographic characteristics of the participants, Efoulan Health District, May 2022 (*n*=46)

| Characteristics | Count (n) | Frequency (%) |
| --- | --- | --- |
| **Health area** |  |  |
| Afanoyoa | 5 | 10.9 |
| Ahala 1 | 3 | 6.5 |
| Ahala 2 | 4 | 8.7 |
| Efoulan | 4 | 8.7 |
| Ngoa-ekelle | 1 | 2.2 |
| Nsimeyong 1 | 8 | 17.4 |
| Nsimeyong 2 | 9 | 19.6 |
| Nsimeyong 3 | 9 | 19.6 |
| Obili | 3 | 6.5 |
| **Age** |  |  |
| 18-24 | 3 | 6.5 |
| 25-34 | 26 | 56.5 |
| 35-44 | 11 | 23.9 |
| 45-49 | 3 | 6.5 |
| 50+ | 3 | 6.5 |
| **Gender** |  |  |
| Female | 39 | 84.8 |
| Male | 7 | 15.2 |
| **Educational level** |  |  |
| Primary | 1 | 2.2 |
| Secondary | 16 | 34.8 |
| Higher | 29 | 63.0 |

Half of the respondents were nurses (58.7%). Half of the participants had already spent more than 2 years in the immunization service and in the health centers where they worked (Table 2).

**Table 2.**
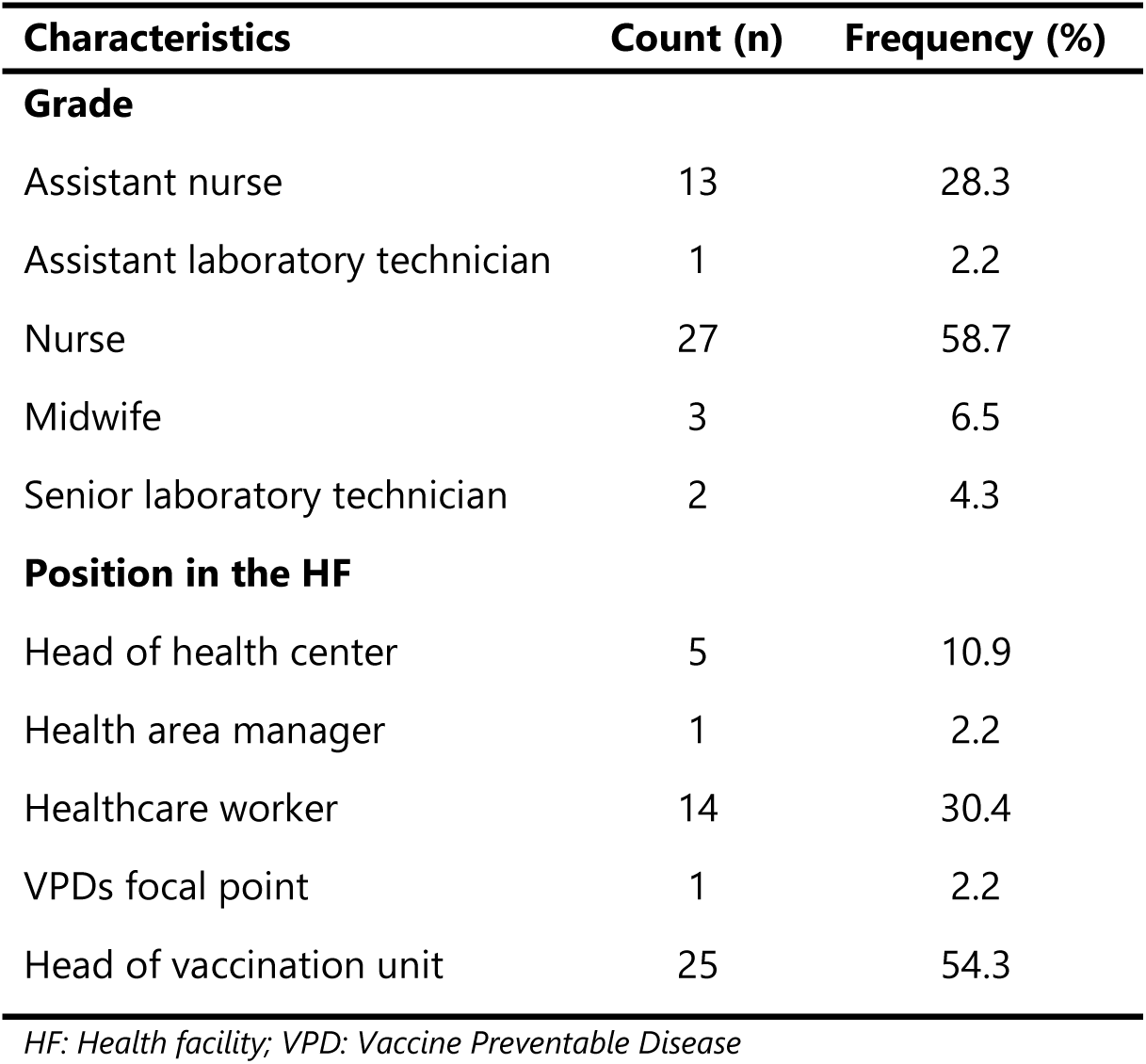
Socioprofessional characteristics of participants, Efoulan Health District, May 2022 (*n*=46)

| Characteristics | Count (n) | Frequency (%) |
| --- | --- | --- |
| **Grade** |  |  |
| Assistant nurse | 13 | 28.3 |
| Assistant laboratory technician | 1 | 2.2 |
| Nurse | 27 | 58.7 |
| Midwife | 3 | 6.5 |
| Senior laboratory technician | 2 | 4.3 |
| **Position in the HF** |  |  |
| Head of health center | 5 | 10.9 |
| Health area manager | 1 | 2.2 |
| Healthcare worker | 14 | 30.4 |
| VPDs focal point | 1 | 2.2 |
| Head of vaccination unit | 25 | 54.3 |

*HF: Health facility; VPD: Vaccine Preventable Disease*

Half of the participants (50%) had already received training on the EPI, and for almost two-thirds of them (65.2%), this training dated back to more than a year. Just over half of the participants reported that they had been supervised as part of their EPI activities, and for most of them (70.4%), this supervision was less than one year old.

### Knowledge of immunization

Fewer than one-third of the participants (26%) identified active artificial immunity as the type of immunity provided by vaccination. More than one-third of the participants (40%) had no knowledge of this topic (Figure 2).

**Fig. 2.**
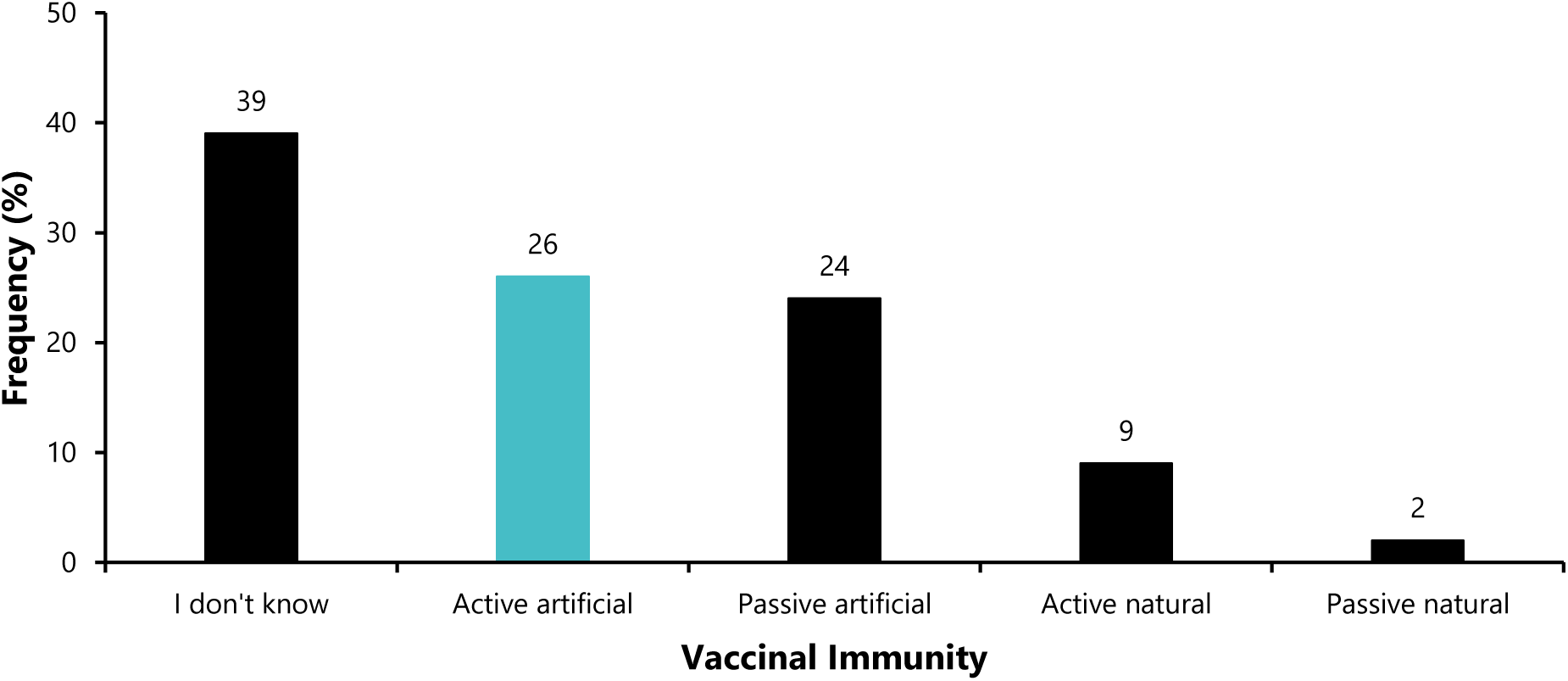
Knowledge of the type of immunity provided by vaccination among healthcare workers, Efoulan Health District, May 2022 (*n*=46)

Almost all participants (93.5%) were unable to name all the diseases the pentavalent vaccine protects against. *Haemophilus influenzae* type b was the antigen they knew least about. Notably, almost one-third of the respondents (30.1%) had no knowledge of the antigens contained in this flagship EPI vaccine (Figure 3).

**Fig. 3.**
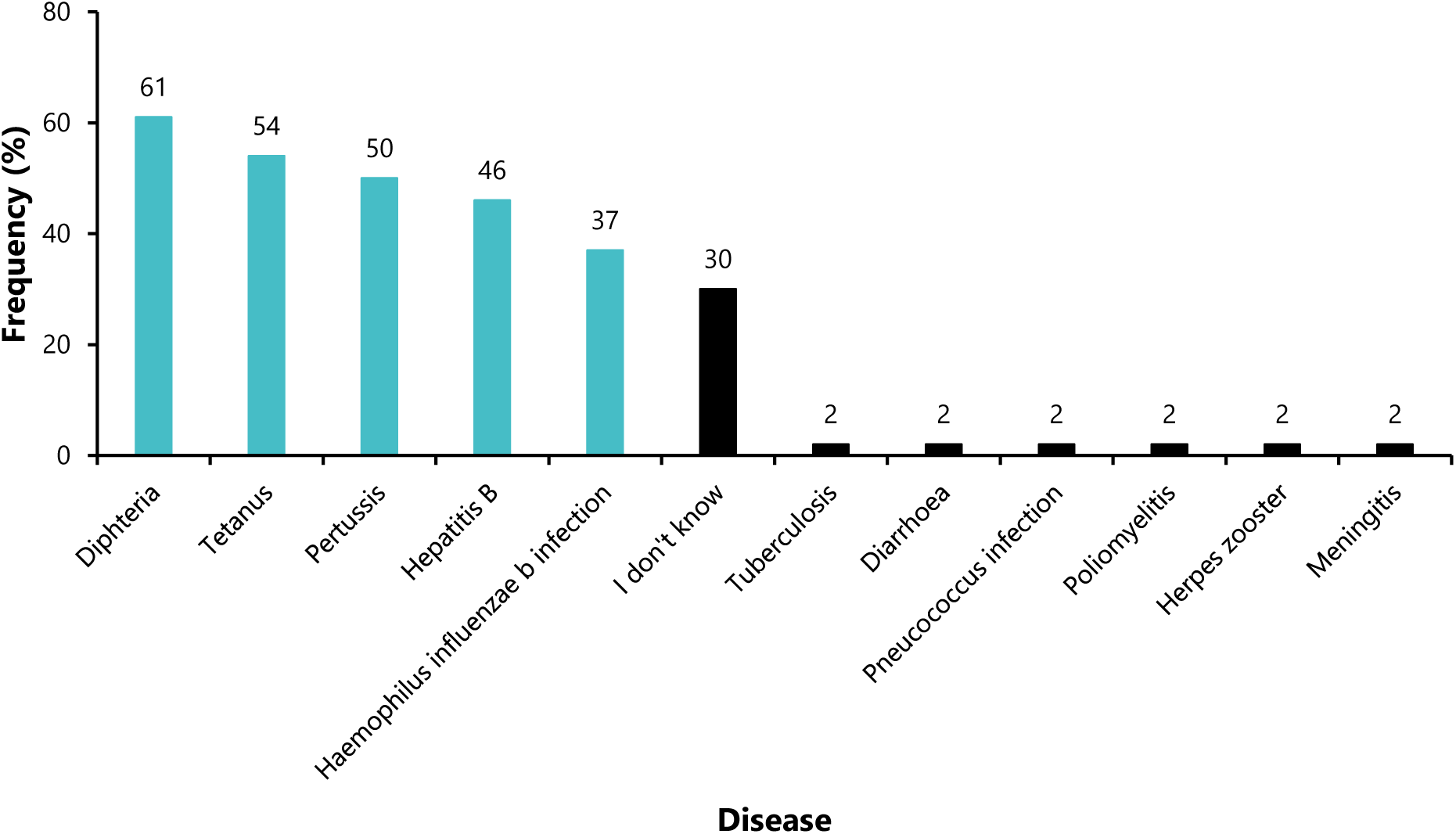
Knowledge of diseases prevented by the pentavalent vaccine among healthcare workers in Efoulan Health District, May 2022 (*n*=46)

More than half of the respondents (59%) were aware of the 15-month booster dose of the measles-rubella (MR)2 vaccine included in the EPI schedule (Figure 4).

**Fig. 4.**
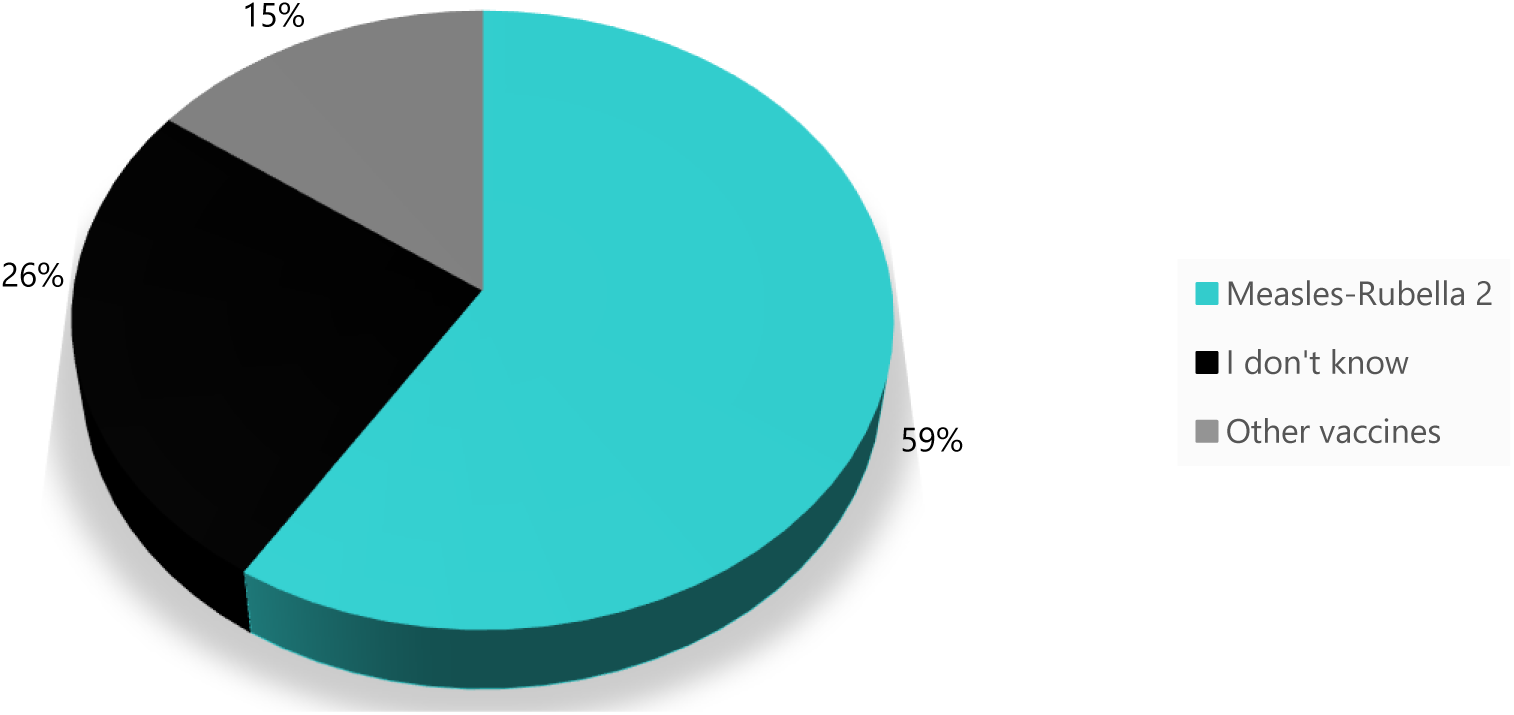
Healthcare workers’ knowledge of the measles-rubella booster at 15 months in the Cameroon EPI schedule, Efoulan Health District, May 2022 (*n*=46)

Half of the respondents were aware of the type of antigen contained in the Bacillus Calmet Guerin (BCG) vaccine (52%) (Figure 5).

**Fig. 5.**
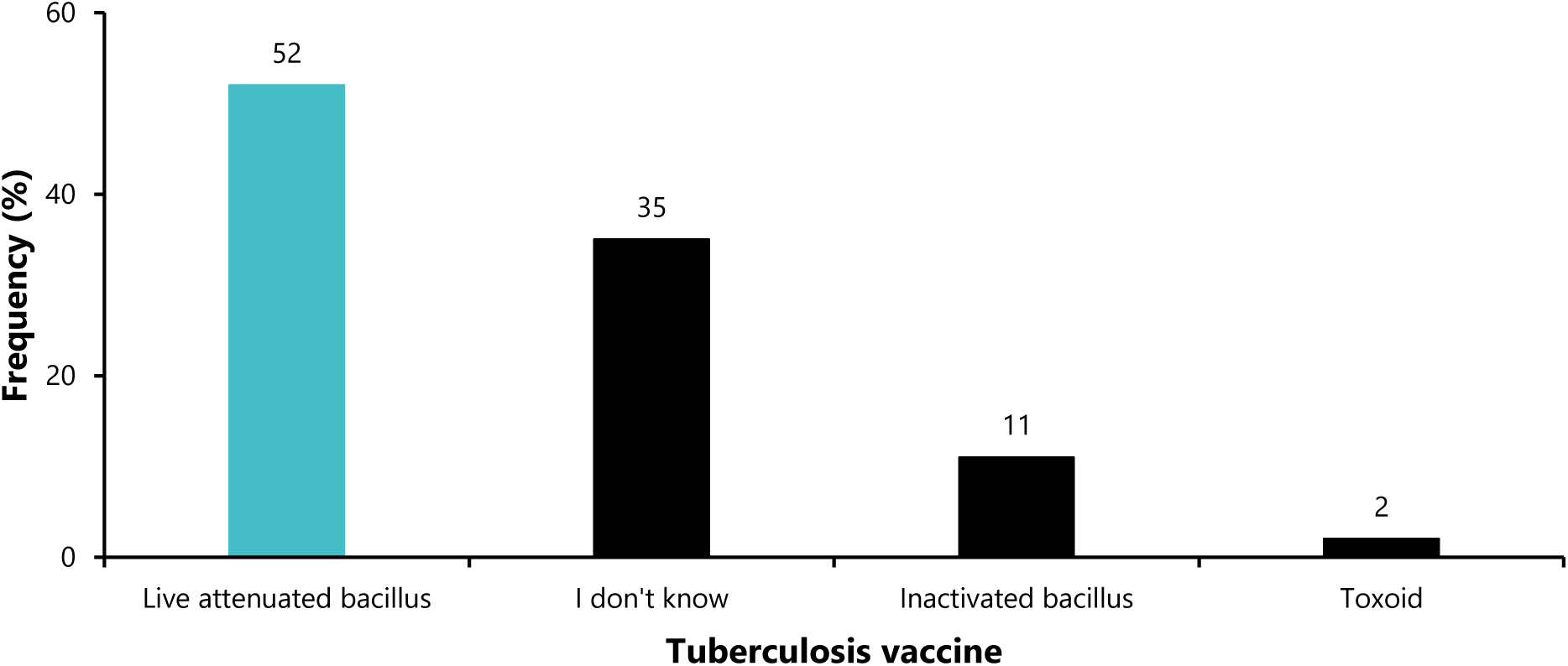
Knowledge of the type of antigen contained in the BCG vaccine among healthcare workers in Efoulan Health District, May 2022.

### Knowledge of priority target populations

A low proportion of participants (6.5%) were aware of the current proportion of surviving children aged 0--11 months expected in Efoulan HDs, which is 2.5%. Slightly more than half of the respondents (59%) had no knowledge of this value, two-thirds of whom (66.7%) had never received EPI training (*p*=0.020). In addition, more than half of the respondents had no knowledge of the expected proportion of pregnant women, which was 2.7% of the total HD population (Table 3).

**Table 3.** Knowledge of priority target proportions among healthcare workers, Efoulan Health District, May 2022 (*n*=46)

| Expected proportion (%) | EPI training received<br>n (%) |  | Total (100%) | *p* value |
| --- | --- | --- | --- | --- |
|  | No<br>*n*=13 | Yes<br>*n*=13 |  |  |
| **Surviving children** |  |  |  |  |
| 2.5 (Yes) | 1 (33) | 2 (67) | 3 | 0.020 |
| 2.7 | 3 (43) | 4 (57) | 7 |  |
| 3.4 | 0 | 6 (100) | 6 |  |
| 4.5 | 1 (33) | 2 (67) | 3 |  |
| I do not know | 18 (67) | 9 (33) | 27 |  |
| **Pregnant women** |  |  |  |  |
| 2.5 | 1 (20) | 4 (80) | 5 | 0.289 |
| 2.7 (Yes) | 1 (20) | 4 (80) | 5 |  |
| 3.4 | 1 (50) | 1 (50) | 2 |  |
| 4.5 | 2 (50) | 2 (50) | 4 |  |
| I don't know | 18 (60) | 12 (40) | 30 |  |

*EPI: Expanded Programme of Immunization*

Most of the participants (87%) did not identify all the vaccines (PCV, Penta and MR) that could be given to an unvaccinated individual one year or more. The most cited vaccine was the MR vaccine (Figure 6).

**Fig. 6.**
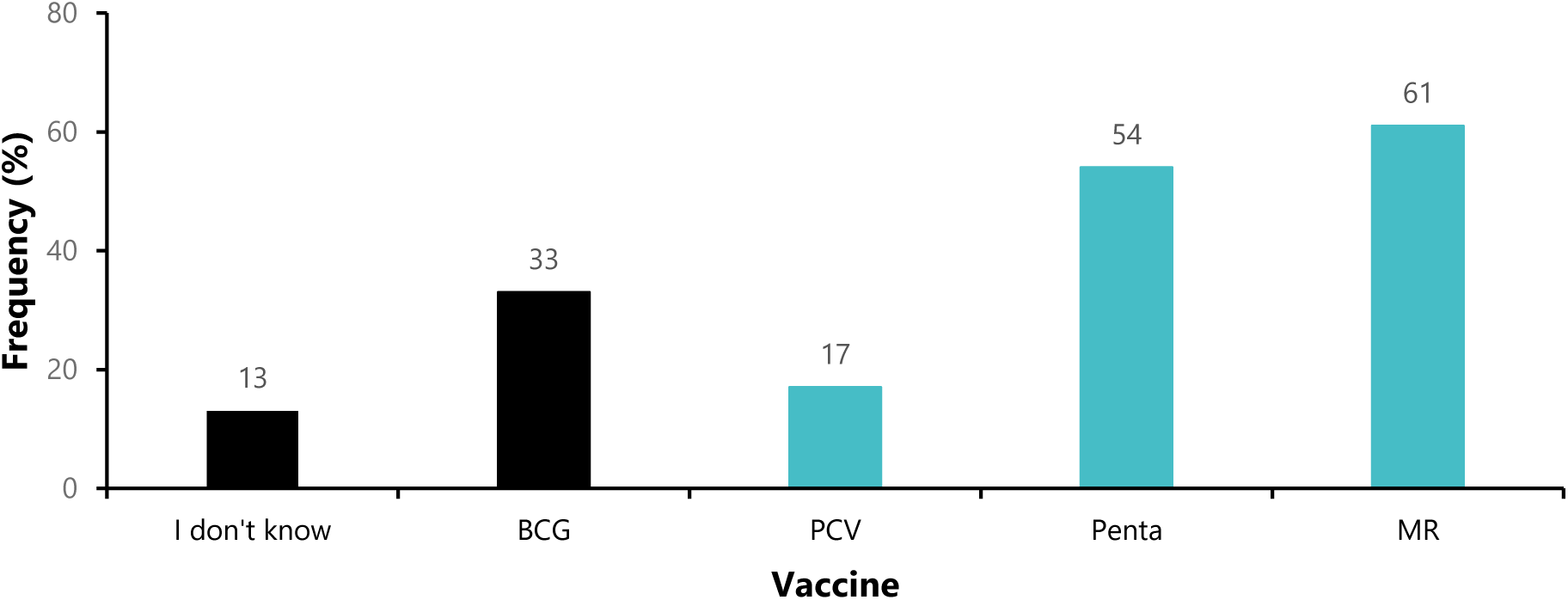
Knowledge of vaccines receivable by an unvaccinated child one year old or older in Efoulan Health District, May 2022 (*n*=46) PCV: Pneumococcal Conjugated Vaccine; BCG: Bacillus Calmet Guerin; MR: Measles-Rubella vaccine; Penta: Pentavalent Vaccine

More than two-thirds of the respondents (72%) had not identified the 6-month MR vaccine as recommended without risk for HIV+ children (Figure 7).

**Fig. 7.**
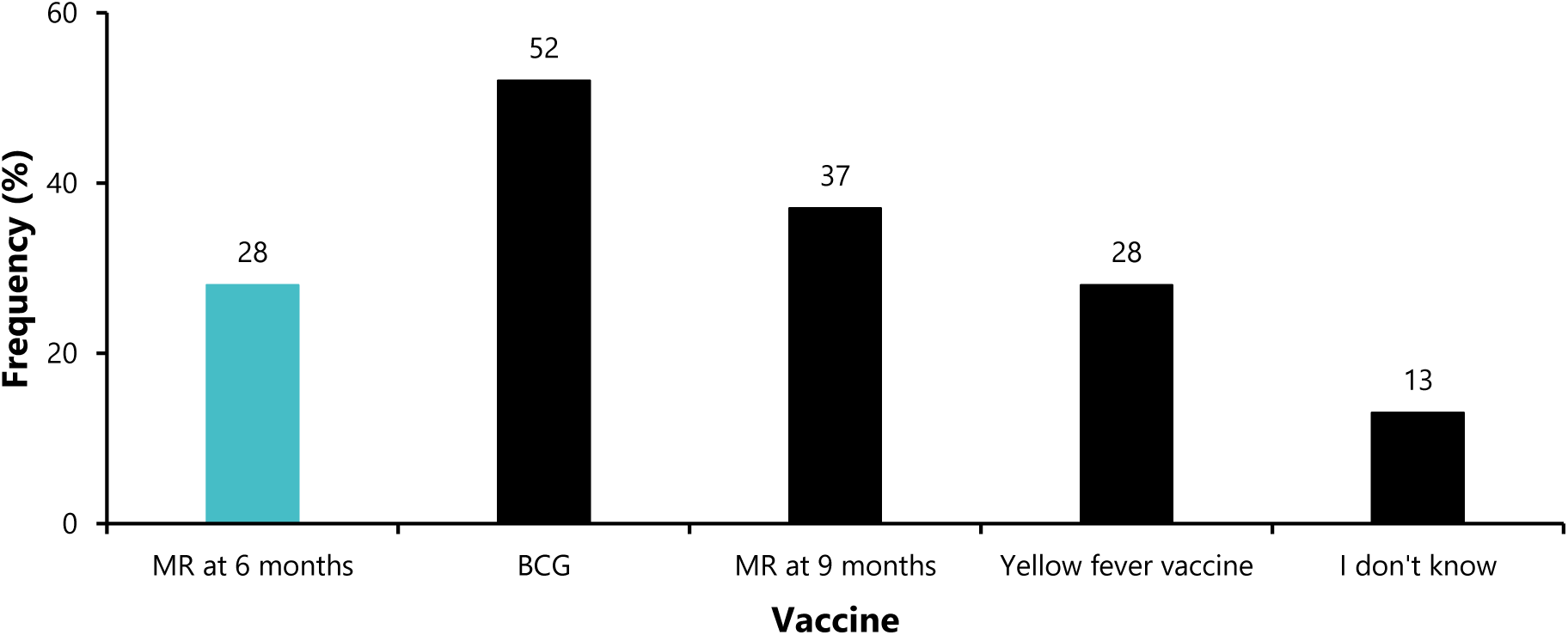
Knowledge of vaccines recommended without risk for HIV+ children among healthcare workers, Efoulan Health District, May 2022 (*n*=46) BCG: Bacillus Calmet Guerin; MR: Measles-Rubella vaccine

Fewer than half of the respondents (44%) identified the optimal 28-day interval between two doses of the EPI vaccine (Figure 8).

**Fig. 8.**
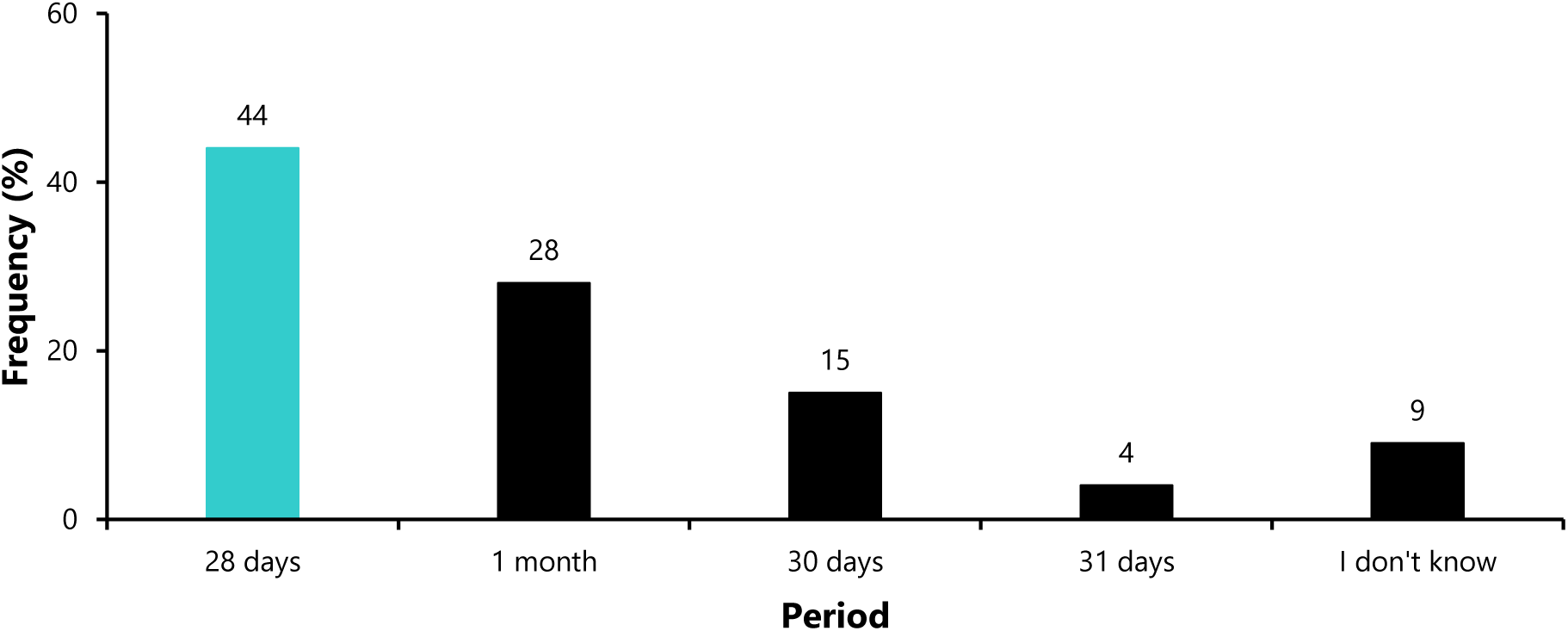
Knowledge of the optimal period between two EPI vaccine doses, Efoulan HD, May 2022 (*n*=46)

Fewer than half of the respondents were aware of the correct angle for intramuscular injection during the vaccination process (43%), with more than a quarter of participants (26%) having no idea about this standard (Figure 9).

**Fig. 9.**
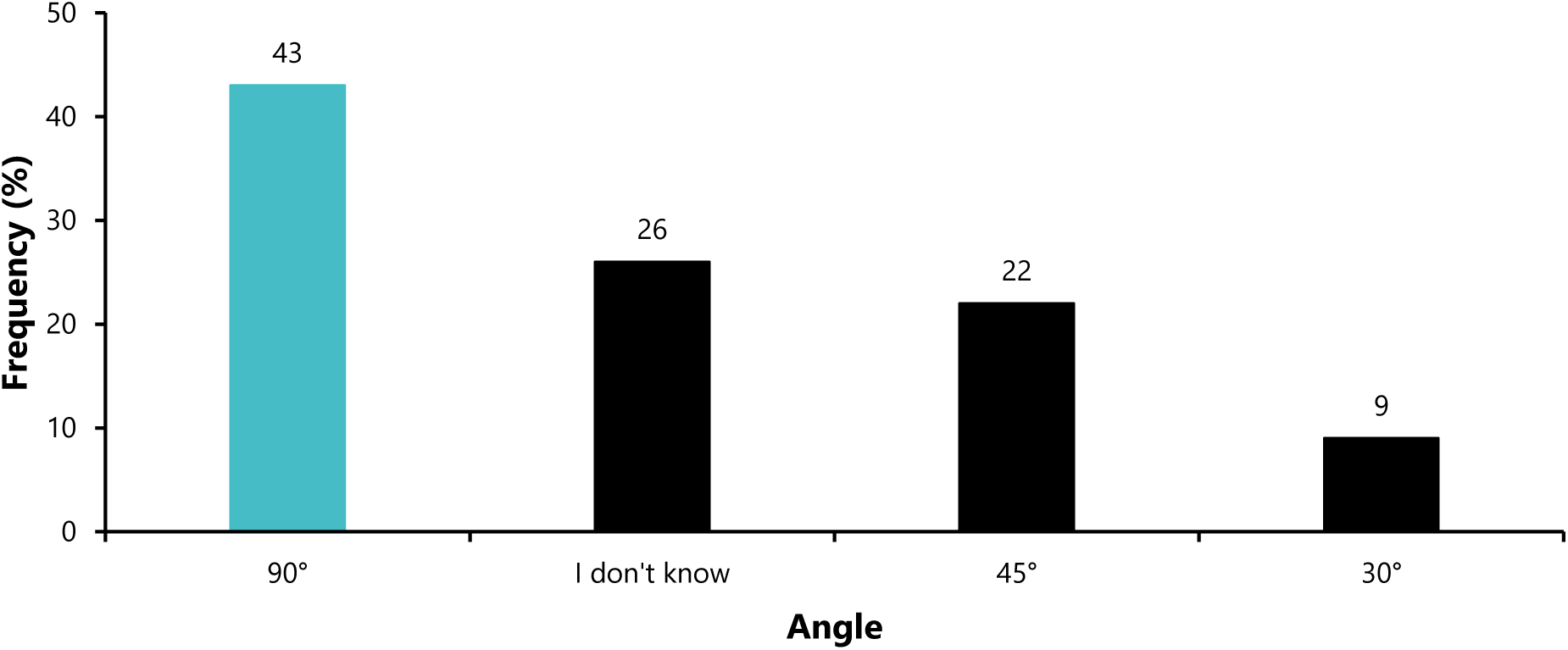
Knowledge of the ideal angle for intramuscular vaccination, Efoulan HD, May 2022 (*n*=46)

Fewer than one-third of HCWs know the distance of 5 km from which the advanced strategy must be implemented. In contrast, more than half of the respondents (52%) did not know the correct distance (Figure 10).

**Fig. 10.**
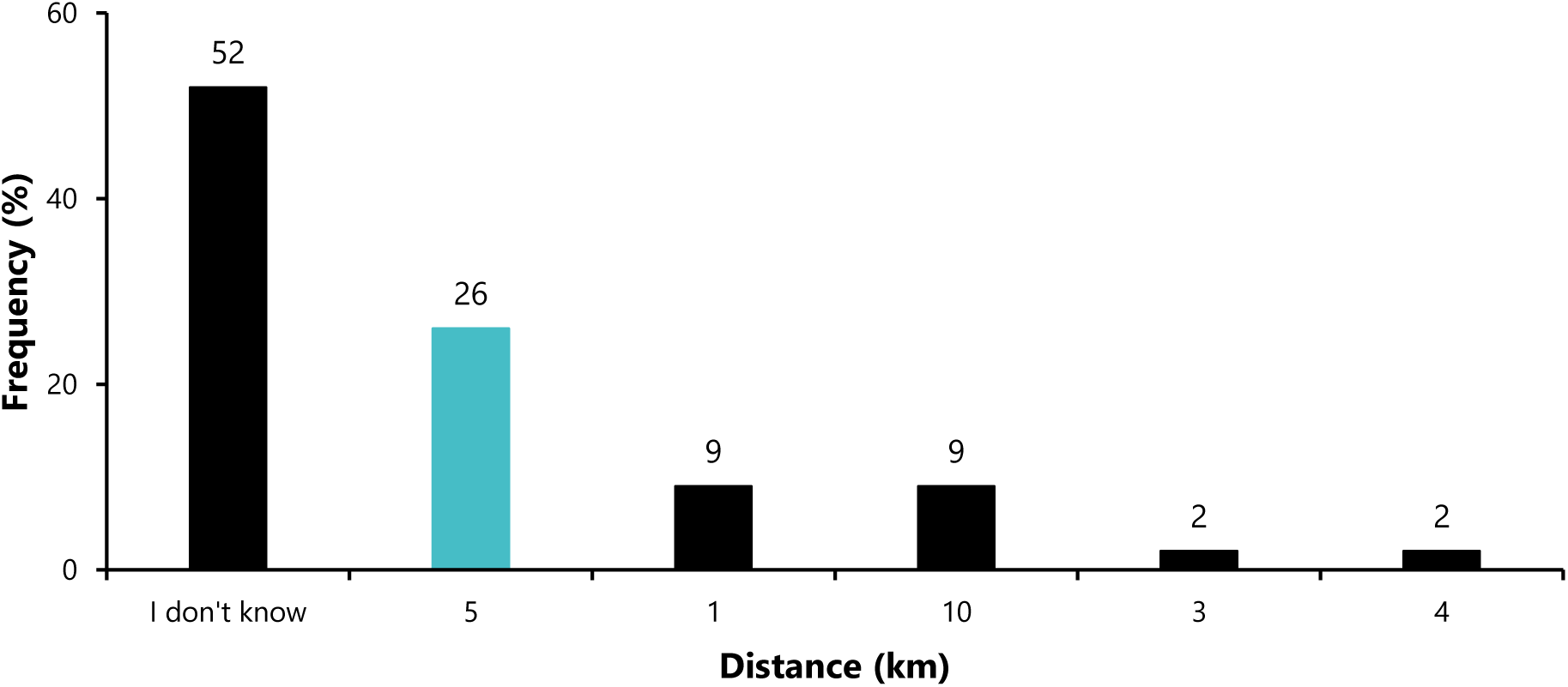
Knowledge of distance requiring organization of advanced vaccination strategy, Efoulan HD, May 2022 (*n*=46)

### Logistic Component of Vaccination

When asked what the EPI tracer antigen was, more than half of the participants correctly identified (41%) penta 3, and more than a quarter (28%) had no idea (Figure 11).

**Fig. 11.**
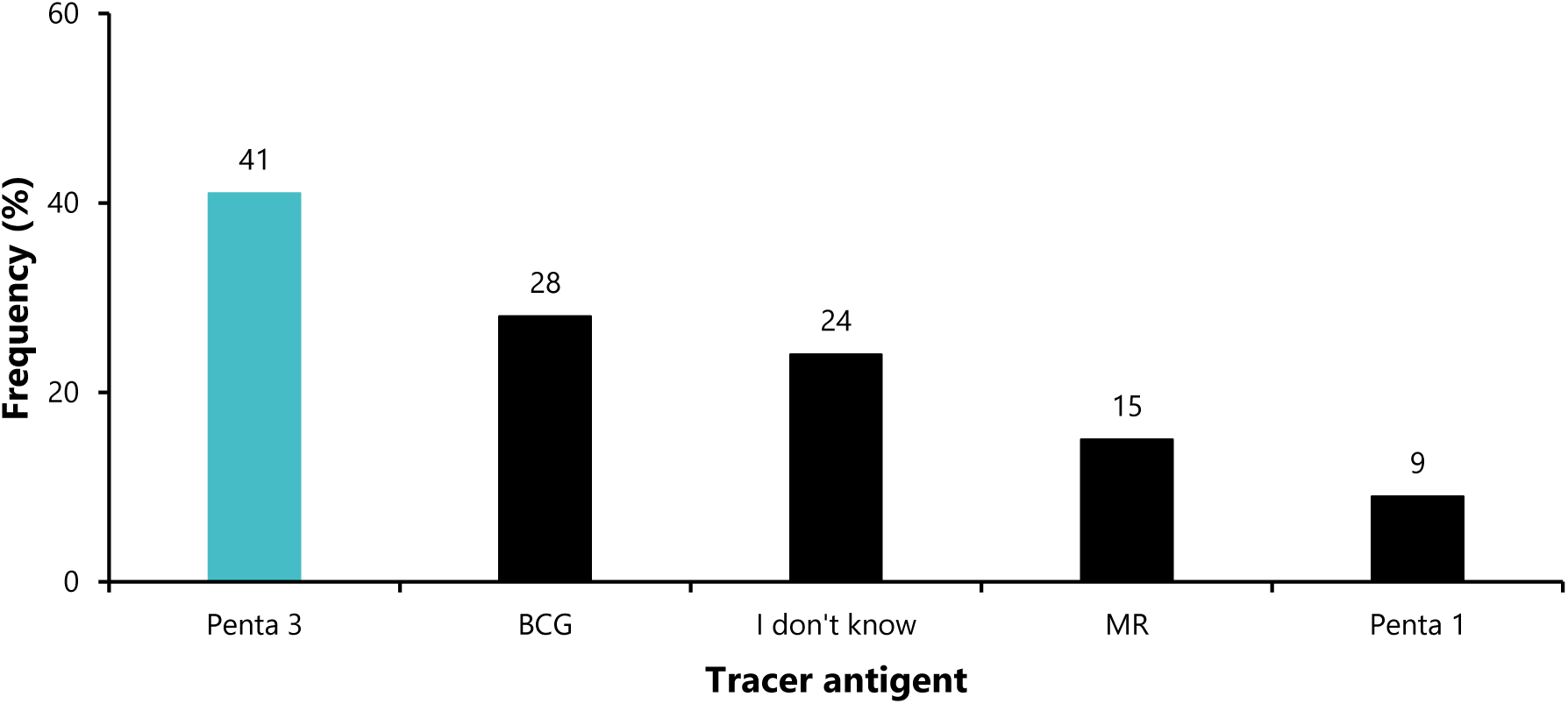
Knowledge of the EPI tracer antigen Efoulan HD, May 2022 (*n*=46) MR: Measles-Rubella; Penta: Pentavalent vaccine; BCG: Bacillus Calmet Guerin

Most of the respondents were not aware of the various components of EPI input management, particularly managing opened nonlyophilized vials (50%), calculating the wastage factor (89%), and calculating vaccine requirements (98%). The participants who reported having received training were significantly more knowledgeable about input management than untrained health workers were (Table 4).

**Table 4.** Knowledge of EPI input management, Efoulan HD, May 2022 (*n*=46)

| Variables | EPI training received |  | Total | *p* value<sup>1</sup> |
| --- | --- | --- | --- | --- |
|  | n (%) |  | (100%) |  |
|  | No<br>*n*=13 | Yes<br>*n*=13 |  |  |
| **Management of opened, nonlyophilized vials** |  |  |  |  |
| Should be discarded after vaccination session | 3 (30) | 7 (70) | 10 | 0.320 |
| Should not be used after 28 days | 11 (48) | 12 (52) | 23 |  |
| Could be used after 28 days | 2 (67) | 1 (33) | 3 |  |
| Did not know how | 7 (70) | 3 (30) | 10 |  |
| **Loss factor** |  |  |  |  |
| No | 1 (11) | 8 (89) | 9 | 0.006 |
| Yes | 1 (20) | 4 (80) | 5 |  |
| Did not know how to calculate | 21 (66) | 11 (34) | 32 |  |
| **Calculation of the requirement dose** |  |  |  |  |
| No | 2 (13) | 13 (87) | 15 | 0.001 |
| Yes | 0 (0) | 1 (100) | 1 |  |
| Did not know how to calculate | 21 (70) | 9 (30) | 30 |  |

<sup>1</sup> Fisher exact test, EPI: Expanded Programme of Immunization

### Global score

For the sixteen questions assessed, the global score ranged from 1/16 (6%) to 11/16 (69%). Half of the participants scored below 25%. The modal score was 3/16 (19%). Almost all the respondents (87%) had poor knowledge of vaccination norms and standards (Table 5).

**Table 5.**
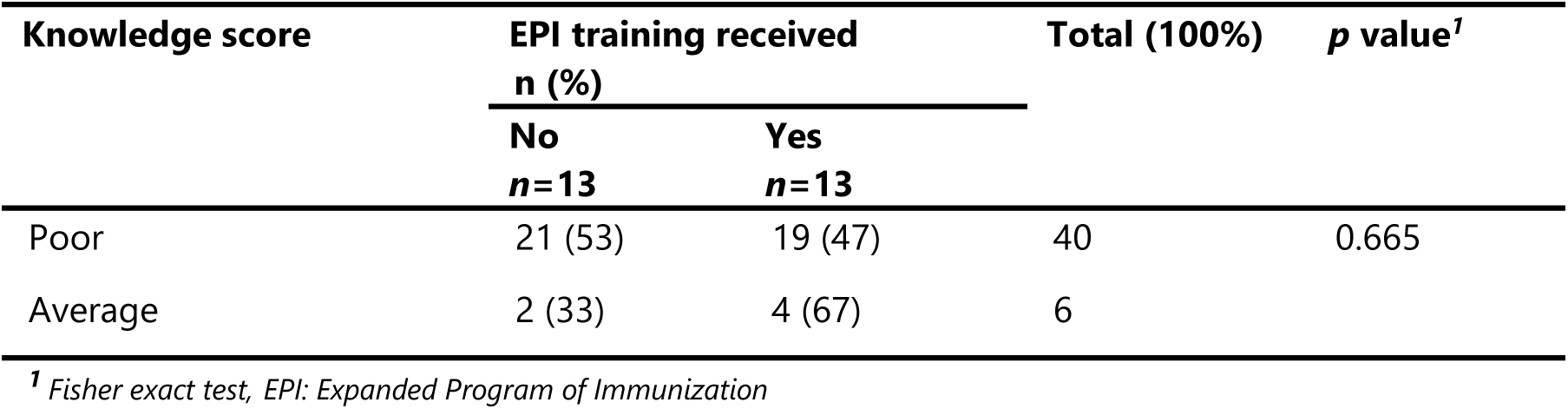
Scores of respondents by previous participation in EPI training, Efoulan Health District, May 2022 (*n*=46)

| Knowledge score | EPI training received |  | Total (100%) | p value<sup>1</sup> |
| --- | --- | --- | --- | --- |
|  | n (%) |  |  |  |
|  | No<br>n=13 | Yes<br>n=13 |  |  |
| Poor | 21 (53) | 19 (47) | 40 | 0.665 |
| Average | 2 (33) | 4 (67) | 6 |  |

<sup>1</sup> Fisher exact test, EPI: Expanded Program of Immunization

### Observance of EPI Norms and Standards

The effective implementation of EPI norms and standards was observed in both public and private HFs of the Efoulan HD identified as problematic (Table 6).

**Table 6.** Health facilities visited in the health areas of the Efoulan Health District, May 2022.

| Health area | Count (n) | Frequency (%) |
| --- | --- | --- |
| Afanoyoa | 2 | 13.3 |
| Ahala 1 | 1 | 6.7 |
| Ahala 2 | 2 | 13.3 |
| Efoulan | 1 | 6.7 |
| Nsimeyong 1 | 1 | 6.7 |
| Nsimeyong 2 | 2 | 13.3 |
| Nsimeyong 3 | 2 | 13.3 |
| Obili | 4 | 26.7 |
| **Total** | **15** | **100** |

HFs vaccinating in the Efoulan HD have several deficiencies in the application of Cameroon’s EPI norms and standards. Most of the assessed HDs did not follow the monthly EPI monitoring curve (87%), did not store vaccines according to standards (80%), or had an EPI case reporting form (93%).

With respect to surveillance, most of the consultants in the HFs visited were not familiar with the definitions of common VPDs (poliomyelitis, yellow fever, measles) (Table 7).

**Table 7.** Health facility compliance with some relevant national EPI guidelines, May 2022 (n= 15)

| Guideline | Frequency<br>n (%) |  |
| --- | --- | --- |
|  | No | Yes |
| **Organization** |  |  |
| Availability of an immunization manager | 0 | 15 (100) |
| Availability of a VPD focal point | 1 (7) | 14 (93) |
| **Implementation and monitoring of the program** |  |  |
| EPI calendar posted | 1 (7) | 14 (93) |
| EPI monitoring curve available | 7 (47) | 8 (53) |
| Monitoring curve plotted and up to date | **13 (87)** | 2 (13) |
| Vaccination registers available and correctly filled in | 5 (33) | 10 (67) |
| Lost to follow-up register available and completed | **10 (67)** | 5 (33) |
| **Cold chain** |  |  |
| Refrigerator available and functional | 1 (7) | 14 (93) |
| Contingency plan posted near the refrigerator | 9 (60) | 6 (40) |
| Fridge temperature record available | 5 (33) | 10 (67) |
| Fridge temperature record updated | **11 (73)** | 4 (27) |
| Fridge temperature measuring device operational | 6 (40) | 9 (60) |
| All EPI antigens available | 9 (60) | 6 (40) |
| Vaccines stored according to standards in the cold chain | **12 (80)** | 3 (20) |
| Tainted vaccines in cold chain | 9 (60) | 6 (40) |
| **Surveillance** |  |  |
| Notification form of VPDs available | **14 (93)** | 1 (7) |
| Definition of measles case known to consultant | 10 (67) | 5 (33) |
| Definition of AFP cases known to the consultant | **12 (80)** | 3 (20) |
| Yellow fever case definition known to the consultant | 9 (60) | 6 (40) |
| VPD cases definitions available and displayed | 4 (26,7) | **11 (73)** |

VPD: Vaccine Preventable Disease; EPI: Expanded Programme of Immunization; AFP: Acute Flaccid Paralysis

## Discussion

The aim of this study was to assess the implementation of vaccination programs in an urban HD in Yaounde. It informs vaccination program implementation and identifies areas for improvement to enhance vaccination coverage and effectiveness in Efoulan HD.

The geographical coverage of the study areas was satisfactory. Most of the HCWs visited had a designated immunization coordinator and a VPD focal point. However, almost half of the HCWs had less than two years of experience in immunization services. This may be due to the high turnover of HCWs between HFs. The retention of skilled HCWs remains critical to the management of human resources for health in Cameroon [13,14]. HCWs, especially those in private HFs, frequently move between HFs in search of better offers and benefits, hindering the effective use of skills acquired through training and supervision.

Significant gaps have been identified in calculating target population sizes, determining loss factors, and estimating vaccine requirements. This situation results in either shortages or surpluses of vaccines in the HF cold chain, which subsequently can lead to expiration and vaccine waste. All this leads to difficulties in stock management and the implementation of a quality vaccination service. This result confirms the findings in Dschang HD in the West Region of Cameroon, where inadequate immunization service delivery was associated with limited knowledge of vaccines and cold chain management by HCWs [15].

Most participants lacked knowledge of the tracer antigen, general vaccine content (e.g., pentavalent vaccine), and safe vaccination techniques (intramuscular injection angle). This inadequacy may affect the ability of HCWs to provide effective education during immunization and to retain mothers for follow-up immunization. An inappropriate route of administration may also increase the risk of adverse events following immunization. As a result, parents may be more reluctant to continue vaccinating their children, leading to an increase in the Penta 3 dropout rate. In addition to an adequate supply of vaccines, the problem of absent or incomplete vaccination could be overcome by improving social mobilization strategies to sensitize mothers and other stakeholders who are reluctant to vaccinate their children, as reported by studies conducted in Cameroon and elsewhere in the world [16–18].

Most HCWs were unaware of the five km distance beyond which advanced strategies must be organized to bring vaccination as close to the population as possible. As a result, advanced vaccination sessions were poorly planned or not organized at all. However, it should also be noted that most HFs in urban HDs are private and that implementing advanced strategies is not part of their culture. This justifies the implementation of supervision of private HFs to sensitize managers and ensure that the benefits of this strategy are well understood and integrated into vaccination planning. Medical evidence has shown that advanced strategies bring immunization services close to the population, reduce the amount of time wasted by parents to get their children vaccinated and increase their adherence to vaccination [19,20].

The observational analysis revealed significant deficiencies in cold chain management and immunization monitoring. Similar trends have been reported in Cameroon and Ethiopia [21,22].

The monitoring curves, although present, were not regularly monitored, and most HFs did not store vaccines properly or monitor temperatures on a daily basis. This is likely due to ignorance or negligence on the part of the people in charge of immunization in these HFs. This situation is attributed to a lack of motivation and poor supervision of HFs by operational and intermediate levels. This result corroborates findings in the North West Health Region in Cameroon [22,23].

Poor understanding of the definitions of cases of VPD and the low level of involvement of HF managers in surveillance, particularly the unavailability of notification forms, could explain the overall underreporting of VPD in the HD.

Overcoming all these challenges will require strong commitment and significant financial mobilization. The shift in healthcare attention during the COVID-19 pandemic can lead to secondary health crises such as outbreaks of infectious diseases like measles. Restrictions due to COVID-19 impede access to HFs for vaccination [17]. During an outbreak such as the COVID-19 pandemic, public health efforts were more focused on controlling the outbreak, resulting in fewer children receiving vaccines. During periods of quarantine, immunization services for all age groups are interrupted, delayed, reorganized, or suspended in many parts of the world [24]. These challenges may have significantly affected the delivery of routine immunization services in the urban HD of Yaounde.

## Limitations

The study has several limitations, particularly the small sample size. A multidistrict study would provide a more comprehensive understanding of the issues in the Central Region. Interpretation of the results should consider the dynamic environment (staff turnover, supervision, and training); therefore, the results may not accurately reflect the current situation in the HD.

## Conclusions

The implementation of routine vaccination remained poor in most of the HFs visited. Significant gaps remained in the EPI-related knowledge of HCWs in HD. Efforts are still needed to increase the level of observance of EPI technical and organizational guidelines to ensure safe and effective immunization of target populations. These efforts include ongoing technical and practical training on EPI guidelines and monitoring to ensure the effective use of the capacity building received by various stakeholders.

## Recommendations

To address the various challenges revealed in this study, we suggest that HDs and regional-level health authorities enhance the training of HCWs, improve the use of vaccination data tools, and strengthen cold chain management and surveillance through planning and implementing the supervision of problematic or priority HFs. They should also develop incentive strategies to retain experienced and trained HCWs.

## Data Availability

All data produced in the present work are contained in the manuscript

## Abbreviations

BCG: Bacillus Calmet Guerin
COVID-19: New Coronavirus Disease
HCW: Healthcare Worker
HD: Health District
HF: Health Facility
HIV: Human Immunodeficiency Virus
MINSANTE: Ministry of Public Health of Cameroon
MR: Measles-Rubella
PCV: Pneumococcal Conjugated Vaccine
Penta: Pentavalent Vaccine
SDG: Sustainable Development Goal
VPD: Vaccine Preventable Disease

## Declarations

### Author contributions

Study design & conception: FZLC; Data collection: FZLC; Data analysis, visualization and interpretation: FZLC; Drafting of original manuscript: FZLC; Critical revision of the manuscript: FZLC, AA, BNA. RKNO, AN, EOG, CSN, AM, GSN, MFE, EABBM, LLEN, CM and FKN; final approval of the manuscript: All authors.

### Ethical Approval Statement

Ethical clearance for the present study was waived by the Faculty of Medicine of Yaounde ethical review board. Administrative approval was obtained from the Chief Medical Officer of Efoulan HD. Additionally, participants were required to provide signed informed consent for their responses to be used for research purposes. The confidentiality, anonymity and autonomy of the research participants were respected throughout the study. All methods were performed in accordance with the relevant guidelines of the Helsinki declaration.

During the field visits, the participants benefited from practical training on the EPI norms and standards. As a result, the various gaps identified in both knowledge and application of the standards were addressed to the best of our ability.

### Consent for publication

Not applicable.

### Availability of data and materials

All data generated or analyzed during this study are included in this published article.

## Acknowledgments

Our gratitude goes to the HCWs of the Efoulan HD who agreed to participate in this study and to the Chief Medical Officer of Efoulan HD, Dr. Ottou Awa Georges Marie, who authorized the conduction of the study in his administrative circumscription.

